# COVID-19 in Multiple Sclerosis Patients and Risk Factors for Severe Infection

**DOI:** 10.1101/2020.05.27.20114827

**Authors:** Farhan Chaudhry, Helena Bulka, Anirudha S. Rathnam, Omar M Said, Jia Lin, Holly Lorigan, Eva Bernitsas, Jacob Rube, Steven J Korzeniewski, Anza B Memon, Phillip D. Levy, Adil Javed, Robert Lisak, Mirela Cerghet

**Affiliations:** Department of Neurology, Henry Ford Health System, Detroit, MI; Wayne State University School of Medicine, Detroit, MI; Department of Neurology, Detroit Medical Center, Detroit, MI; Department of Neurology, University of Chicago, Chicago, Illinois, USA

## Abstract

**Importance:** Multiple sclerosis patients have been considered a higher-risk population for COVID-19 due to the high prevalence of disability and disease-modifying therapy use; however, no study has identified clinical characteristics of multiple sclerosis associated with worse COVID-19 outcomes.

**Objective:** To evaluate the clinical characteristics of multiple sclerosis, including staging, degree of disability, and disease-modifying therapy use that are associated with worse outcomes from COVID-19.

**Design:** Prospective cohort study looking at the outcomes of multiple sclerosis patients with COVID-19 between March 1^st^ and May 18^th^ 2020.

**Setting:** This is a multicenter study of three distinct hospital systems within the U.S.

**Participants:** The study included 40 consecutive patients with nasopharyngeal/oropharyngeal PCR-confirmed COVID-19 infection.

**Exposures:** Multiple sclerosis staging, severe disability (based on baseline-extended disability status scale equal to or greater than 6.0) and disease-modifying therapy.

**Main Outcomes and Measure:** Severity of COVID-19 infection was based on hospital course, where a mild course was defined as the patient not requiring hospital admission, moderate severity was defined as the patient requiring hospital admission to the general floor only, and most severe was defined as requiring intensive care unit admission and/or death.

**Results:** For the 40 patients, the median age was 52(45.5-61) years, 16/40(40%) were male, and 21/40(52.5%) were African American. 19/40(47.5%) had mild courses, 15/40(37.5%) had moderate courses, and 6/40(15%) had severe courses. Patients with moderate and severe courses were significantly older than those with a mild course (57[50-63] years old and 66[58.8-69.5] years old vs 48[40-51.5] years old, P=0.0121, P=0.0373). There was differing prevalence of progressive multiple sclerosis staging in those with more severe courses (severe:2/6[33.3%]primary-progressing and 0/6[0%]secondary-progressing, moderate:1/14[7.14%] and 5/14[35.7%] vs mild:0/19[0%] and 1/19[5.26%], P=0.0075, 1 unknown). Significant disability was found in 1/19(5.26%) mild course-patients, but was in 9/15(60%, P=0.00435) of moderate course-patients and 2/6(33.3%, P=0.200) of severe course-patients. Disease-modifying therapy prevalence did not differ among courses (mild:17/19[89.5%], moderate:12/15[80%] and severe:3/6[50%], P=0.123).

**Conclusions and Relevance:** Multiple sclerosis patients with more severe COVID-19 courses tended to be older, were more likely to suffer from progressive staging, and had a higher degree of disability. However, disease-modifying therapy use was not different among courses.

## Introduction

Patients with multiple sclerosis (MS) are four times more likely to succumb to serious infection when compared to the general population, making them potentially at higher-risk from the ongoing novel coronavirus disease (COVID-19).^1^ This may be because a significant number of MS patients have a high level of disability and are on immunosuppressive disease-modifying therapies (DMTs). Most information regarding the association of MS characteristics and DMT- usage with COVID-19 comes from case-reports and case-series surveys.^2,3^ In this prospective cohort study we looked at the MS disease characteristics with differing COVID-19 severity.

## Methods

### Study Design

We enrolled 40 consecutive MS patients with confirmed COVID-19 via nasopharyngeal/oropharyngeal-PCR from 3 different hospital systems in the U.S, Henry Ford Hospital Systems (HFHS) in Detroit, MI, Detroit Medical Center (DMC) in Detroit MI, and University of Chicago (UOC) in Chicago, IL (28 at HFHS, 4 at DMC and 8 at UOC) between March 1^st^ and May 18^th^. Study was approved by each institution review board (IRB).

### Data Collection

We obtained information regarding age, race, sex, smoking status, MS stage (Primary Progressive MS, PPMS; Relapsing-Remitting MS, RRMS; Secondary Progressive MS, SPMS), duration of MS, DMT, and comorbidities. Extended disability status scale (EDSS) was used for disability assessment with ≥ 6.0 indicating severe disability.^4,5^ Symptoms and labs at presentation were also included in the analysis. Days of COVID-19 symptoms was a patient-reported metric.

### Outcome

Clinical severity of COVID-19 was based on hospital course: mild course was defined as not requiring hospital admission, moderate course was defined as requiring admission to general floor only, and most severe course was defined as requiring intensive care unit (ICU) admission and/or death.^6^

### Statistical Analysis

R-version 3.6.3 was used for statistical analysis. Continuous variables were reported as median with interquartile ranges (median[IQR]). Categorical variables were reported as counts (percentages). Lab data was expressed as frequency of elevated labs using cut off values (Supplement Table 1) and median[IQR]. Continuous variables were compared using Kruskal-Wallis-H test with post hoc Dunn’s test for pairwise-comparisons. Due to small sample size, proportions of categorical variables were compared using Chi-squared test with Monte-Carlo simulation with 2000 replicates.^7^ Then Fisher multiple-pairwise tests were used for post hoc comparisons with Benjamini-Hochberg adjustments.^8^ Differences among COVID-19 courses and pair-wise comparisons were considered statistically significant if P-value or adjusted P-value was less than 0.05.

## Results

20/40(50%) patients were admitted, of which 5/20(25%) patients required ICU admission. 4/20(20%) were ventilated, 3/20(15%) died, and 1/20(5%) made a full recovery. One ICU patient only required continuous positive airway pressure and made a full recovery. One patient died on arrival at the emergency department, a total of 4/40(10%) of patients, while all other patients recovered (36/40 [90%]).

19/40(47.5%) had mild courses, 15/40(37.5%) had moderate courses, and 6(15%) had severe courses. Moderate (57[50-63 years old) and severe (66[58.8-69.5] course-patients were significantly older than mild (48[40-51.5]) course-patients (P=0.0121, P=0.0373 respectively). Moderate course-patients were more likely to be smokers compared to mild course-patients (5/14[35.7%] vs 0/19[0%], P=0.0253).

There was differing prevalence of progressive forms of MS among courses of COVID-19 (severe: 2/6[33.3%]PPMS and 0/6[0%]SPMS; moderate:1/14[7.14%]PPMS and 5/14[35.7%]SPMS vs mild: 0/19[0%]PPMS and 1/19[5.26%]SPMS, P=0.0075, 1 unknown). Moderate course-patients tended to have MS longer than patients with mild and severe courses (16[11.5-27] years vs 11 [6-14.5] years and 5[3-10] years, P=0.0480, P=0.283).(Table 1a)

**Table 1:** Patient Characteristics. Characteristics of all patients. a) Patient demographic data, b) Comorbidities, c) Initial presentation, d) Treatment, e) Outcomes. DMT= Disease Modifying Therapies (Details on MS drugs shown in Supplementary Table 1), PPMS=Primary progressive MS, RRMS= Relapse-remitting MS, SPMS=Secondary progressive MS. EDSS= Expanded Disability Status Scale. BMI= Body Mass Index. All continuous variables are expressed with median and interquartile range as median (IQR). All frequencies are presented as fraction of counts over number of available counts (Percentage). Lab data is expressed as frequency of elevated labs (%); median (IQR).(Reference values shown in Supplementary Table 1) Lactate Dehydrogenase(LDH), C-reactive protein (CRP), Creatine Phosphokinase (CPK), Liver Function Test (LFT). *= statistically significant difference compared with mild course, ^^^=statistically significant difference compared with moderate course.

|  | All | Mild | Moderate | Severe | P-value |
| --- | --- | --- | --- | --- | --- |
| n | 40 | 19 | 15 | 6 |  |
| **a) Demographics and MS characteristics** |  |  |  |  |  |
| Age in years(IQR) | 52 (45.5-61) | 48 (40-51.5) | 57 (50-63)* | 66 (58.8-69.5)* | 0.0013 |
| African American | 21/40 (52.5%) | 7/19 (36.8%) | 9/15 (60%) | 5/6 (83.3%) | 0.102 |
| Male | 16/40 (40%) | 5/19 (26.3%) | 8/15 (53.3%) | 3/6 (50%) | 0.258 |
| BMI (IQR) | 30.6 (25.5-35.9) | 30.7 (27-36.4) | 28.4 (23.1-33.8) | 36.1(34.2-38.8) | 0.142 |
| Smoker | 6/39 (15.4%) 1 not specified | 0/19 (0%) | 5/14 (35.7%)* | 1/6 (16.7%) | 0.0145 |
| *Stage of MS* |  |  |  |  |  |
| PPMS | 3/39 (7.69%), 1 not specified | 0/19 (0%) | 1/14 (7.14%)* | 2/6 (33.3%) | 0.0075 |
| RRMS | 30/39 (76.9%) | 18/19 (94.7%) | 8/14 (57%)* | 4/6 (66.7%) |  |
| SPMS | 6/39 (15.4%) | 1/19 (5.26%) | 5/14 (35.7%)* | 0/6 (0%) |  |
| Years with MS(IQR) | 12 (6.5-19) | 11 (6-14.5) | 16 (11.5-27)* | 5 (3-10)^ | 0.0166 |
| EDSS ≥6.0 | 12/40 (30%) | 1/19 (5.26%) | 9/15 (60%)* | 2/6 (33.3%) | 0.0045 |
| On DMT | 32/40 (80%) | 17/19 (89.5%) | 12/15 (80%) | 3/6 (50%) | 0.123 |
| Nursing Home | 8/40 (20%) | 0/19 (0%) | 7/15 (46.7%)* | 1/6 (16.7%) | 0.0015 |
| Healthcare Worker | 6/40 (15%) | 5/19 (26.3%) | 1/15 (6.67%) | 0/6 (0%) | 0.132 |
| Baseline Absolute Lymphocyte Count(IQR) | 1.6 (0.9-1.8) | 1.7 (1.35-1.72) | 1.4(0.925-1.85) | 1.6(1.2-2.8) | 0.908 |
| **b) Comorbidities** |  |  |  |  |  |
| Hypertension | 16/40 (40%) | 3/19 (15.8%) | 10/15 (66.7%)* | 3/6 (50%) | 0.0085 |
| Hyperlipidemia | 7/40 (17.5%) | 1/19 (5.26%) | 4/15 (26.7%) | 2/6 (33.3%) | 0.172 |
| Diabetes | 6/40(15%) | 1/19 (5.26%) | 4/15 (26.7%) | 4/6 (66.7%)* | 0.005 |
| Asthma/COPD | 2/40 (5%) | 0/19 (0%) | 1/15 (6.67%) | 1/6 (16.7%) | 0.277 |
| **c) Presentation** |  |  |  |  |  |
| Fever | 26/40 (65%) | 10/19 (52.6%) | 11/15 (73.3%) | 5/6 (83.3%) | 0.331 |
| Cough | 26/40 (65%) | 16/19 (84.2%) | 6/15 (40%)* | 4/6 (66.7%) | 0.0275 |
| Myalgia | 14/40 (35%) | 9/19 (47.4%) | 3/15 (20%) | 2/6 (33.3%) | 0.302 |
| Shortness of Breath | 20/40 (50%) | 7/19 (36.8%) | 8/15 (53.3%) | 5/6 (83.3%) | 0.148 |
| Diarrhea | 4/40 (10%) | 1/19 (5.26%) | 3/15 (20%) | 0/6 (0%) | 0.275 |
| Sore Throat | 4/40 (10%) | 3/19 (15.8%) | 0/15 (0%) | 1/6 (16.7%) | 0.366 |
| Headache | 7/40 (17.5%) | 3/19 (15.8%) | 4/15 (26.7%) | 0/6 (0%) | 0.42 |
| Altered Mental Status | 4/40 (10%) | 0/19 (0%) | 4/15 (26.7%) | 0/6 (0%) | 0.0185 |
| Require Chest X-Ray | 23/40(57.5%) | 2/19(10.5%) | 15/15(100%)* | 6/6(100%)* | 0.0005 |
| Chest X-Ray Abnormality | 17/23 (73.9%) | 2/2 (100%) | 9/15 (60%) | 6/6 (100%) | 0.132 |
| Elevated Troponin | 3/14 (21.4%), 26 labs not obtained | 0/1 (0%) | 2/12 (16.7%) | ¼ (25%) | 0.371 |
| Elevated LDH; median(IQR) IU/L | 15/16 (93.8%), 24 labs not obtained; 357(254-433) | 0/0 | 11/12 (91.7%); 312(245-394) | 4/4 (100%);598 (402-807) | 0.0452 |
| Elevated Procalcitonin; median(IQR) ng/mL | 9/13 (69.2%), 27 labs not obtained; 0.31 (0.02-0.57) | 0/0 | 7/10 (70%); 0.295 (0.08-0.507) | 2/3 (66.7%);1.07 (0.535-6.07) | 0.307 |
| Elevated Ferritin; median(IQR) ng/mL | 12/16 (75%), 24 labs not obtained;703.5(288-1107) | 0/0 | 9/12 (75%); 494 (195-1030) | 3/4 (75%);980 (675-1218) | 0.332 |
| Elevated D-Dimer; median(IQR) ug/mL | 14/16 (87.5%), 24 labs not obtained; 2.14 (0.988-3.32) | 0/0 | 9/11 (81.8%); 2.15(0.985-2.84) | 5/5 (100%);2.13 (1.91-5.5) | 0.461 |
| Elevated CRP;median(IQR) mg/dL | 16/16 (100%) 24 labs not obtained: 9 (5-13) | 0/0 | 12/12 (100%); 9 (4-13) | 4/4 (100%);9.5 (7.25-12) | 0.734 |
| Elevated CPK; median(IQR) IU/L | 7/15 (46.7%), 25 labs not obtained; 290 (85-442.5) | 0/0 | 6/11 (54.5%); 94(79.5-442) | 2/4 (50%);308 (238-3296) | 0.514 |
| Elevated LFT; median(IQR) IU/L | 10/20 (50%), 20 labs not obtained; 24.5 (19.5-40.8) ALT, 30 (22.75- | 0/1 (100%);21ALT, 17AST | 8/14 (57.1%); 24 (18.5-37)ALT, 34 (24.5- | 2/5 (40%);32 (25-70) ALT,28 (22-121)AST | 0.638 |
|  | 49.3) AST |  | 48)AST |  |  |
| White Blood Cell Count (IQR) K/uL | 6.5 (5.20-8.3) | 4.3 | 5.6 (5.2-9.35) | 7.8 (7.1-8.2) | 0.352 |
| Neutrophil%(IQR) | 79 (65-86) | 52 | 79 (64.5-85.5) | 83 (80-88) | 0.163 |
| Lymphocyte%(IQR) | 12 (9-15) | 38 | 12 (9-18) | 13 (7-14) | 0.313 |
| **d) Treatment specifically for COVID-19** |  |  |  |  |  |
| Given Treatment | 21/40 (52.5%) | 4/19 (21.1%) | 12/15 (80%)* | 5/6 (83.3%)* | 0.001 |
| Hydroxychloroquine | 14/40 (35%) | 1/19 (5.26%) | 8/15 (53.3%)* | 5/6 (83.3%)* | 0.001 |
| Antibiotic | 13/40 (32.5%) | 3/19 (15.8%) | 6/15 (40%) | 4/6 (66.7%) | 0.048 |
| Methylprednisone | 10/40 (25% %) | 0/19 (0%) | 6/15 (40%)* | 4/6 (66.7%)* | 0.002 |
| **e) Outcome** |  |  |  |  |  |
| Days Admitted(IQR) | 8 (4-14) | NA | 4 (4-9.5) | 15.5 (14.2-16.8) | 0.00601 |
| Days with COVID-19 Symptoms(IQR) | 16 (13-20.3) | 16 (14-21) | 14 (4-18.5) | 16.5 (14.2-21.8) | 0.304 |
| Require Ventilator | 4/40 (10%) | NA | NA | 4/6 (66.7%) |  |
| Death | 4/40 (10%) | NA | NA | 4/6 (66.7%) |  |

EDSS was ≥ 6.0 in 1/19(5.26%) with the mild course, significantly less than that with moderate courses (9/15[60%], P=0.00435). This was also less than that seen in severe course-patients, though not statistically significant (2/6[33.3%], P=0.200). More patients with moderate courses were residents of nursing homes compared to those with mild courses (7/15 [46.7%] vs 0/19[0%], P=0.0036).

There was a relatively higher prevalence of DMT use in patients with mild and moderate courses compared to those with severe courses, although not found to be statistically significant across groups; 17/19[89.5%] and 12/15[80%] vs. 3/6[50%], P=0.123). Percentages of different types of DMT appeared to be similar across courses. (Figure 1)

**Figure 1:**
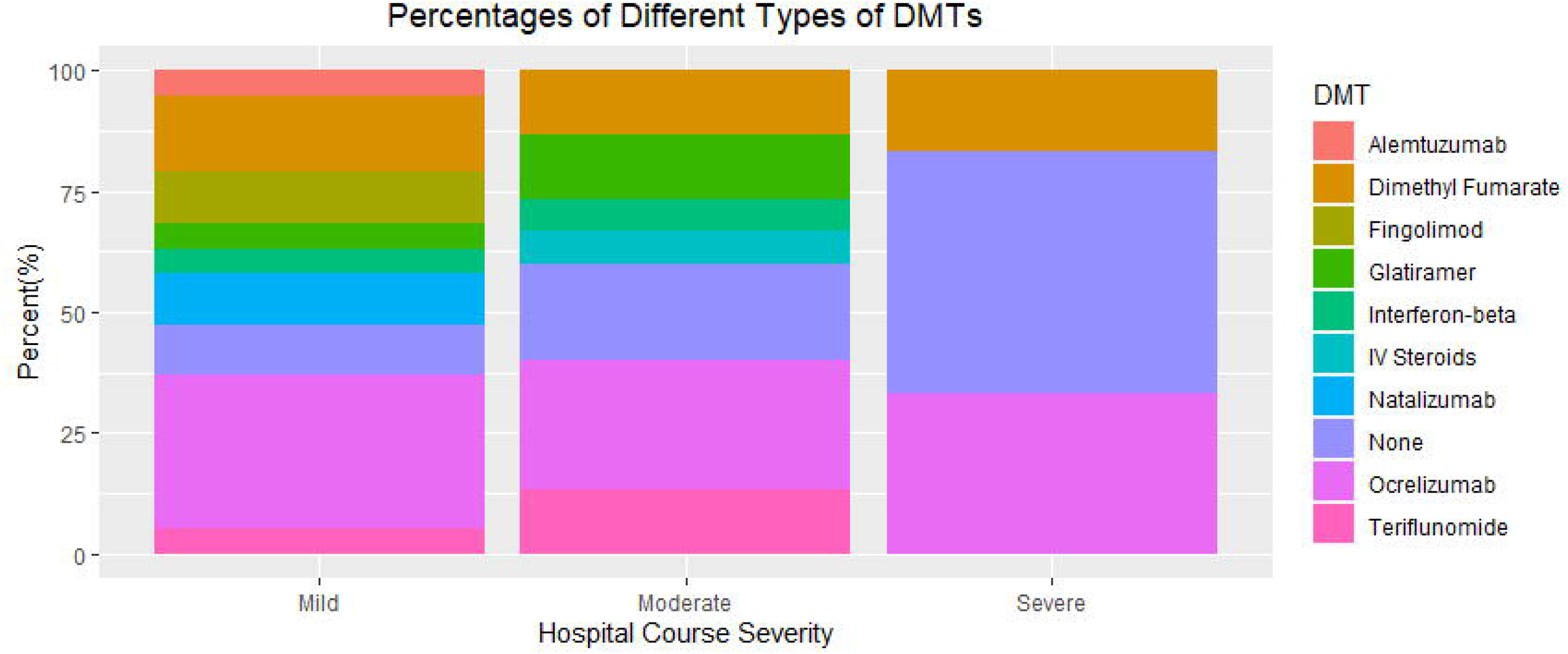
Percentages of Different Types of DMTs. A bar graph showing the relative percentages of different types of DMTs for each hospital course.

There were more patients with moderate courses who had hypertension compared to that seen in those with mild courses (10/15[66.7%] vs 3/19[15.8%], P=0.0127). There were more patients with severe courses who had diabetes compared to that seen in those with mild courses (4/6[66.7%] vs 1/19[5.26%], P=0.005).(Table 1b)

Overall, initial presenting symptoms were similar among each course of COVID-19. (Table 1c) All moderate and severe courses, as expected, had chest X-ray obtained compared to only 2/19(10.5%) patients with mild courses (P=0.0005). Patients with severe courses had quantifiably higher levels of lactate dehydrogenase (LDH, 598[402-807]IU/L vs 312[245-394]IU/L, P=0.0452). Patients with moderate and severe courses were more likely to be treated for COVID-19 compared to those with mild courses (12/15[80%] and 5/6[83.3%] vs4/19[21.1%], P=0.00429, P=0.0178).(Table 1c)

All admitted patients were not given DMTs during admission. Duration of COVID-19 symptoms was similar among all courses(P=0.304). Patients with severe courses were more likely to be admitted longer compared to those with moderate courses (15.5[14.2-16.8] days vs 4[4-9.5] days, P=0.00601).(Table 1e)

## Discussion

MS is a heterogeneous disease as PPMS and SPMS are associated with a later age of onset when compared to RRMS, thus patients tend to be older with more disablities.^9,10^ Older age, certain comorbidities and living in nursing homes are significant risk factors for COVID-19.^11^ Indeed, MS patients with moderate-severe courses were more likely to have progressive disease, comorbidities (smoking, hypertension, and diabetes), and EDSS scores ≥ 6.0. These patients also tended to be older and frequently resided in nursing homes.

It is known that certain DMTs have higher infectious risk compared to others; therefore, it has been a concern that cell depleting DMTs would be associated with higher COVID-19 risk.^12^ However, in our cohort, there doesn’t appear to be bias regarding certain DMTs in moderate-severe courses. In fact, it appears that more patients with moderate and severe courses were not on any DMT compared to that seen in those with mild courses. DMTs have been hypothesized to attenuate the cytokine-storm response in COVID-19, though this is still purely speculative.^12^

Overall, usage of DMTs do not necessarily appear to be associated with more severe COVID-19-courses. These findings correspond to those in ongoing North American COVID-19 MS survey database (covidms.org) and Italian cohort surveyed by Sormani el al. where patients who died tended to be older, have PPMS or SPMS, and have significant disability and comorbidities, but were less likely to be on DMT.^3,13^

As of 5/25/2020, the Detroit Health Department has reported a 12.3% citywide COVID-19 mortality rate among laboratory confirmed cases.^14^ Although, we had a small sample size, our MS cohort reported a similar mortality rate and was like that reported by Sormani et al. in confirmed-COVID-19 positive MS patients (5/57[8.78%]).^3^ Therefore, we can assume our mortality rate could be consistent with a larger sample size and similar to that of the general population.

A significant limitation to our study is the relatively small sample size collected; therefore risk-stratification adjusting for confounders was not possible. Larger studies are therefore required in order to improve for confounding variables. Also, there was a significant discrepancy in sample-size for each course, as there were only 6 patients with severe disease course. Therefore, significant differences may not have been detected in post hoc comparisons with this course. Regardless, we showed that certain MS-characteristics were more frequent in more COVID-19 severe courses.

## Data Availability

Due to the sensitivity of protected health information, de-identified data will only be available upon request to corresponding author.

## Acknowledgement

We dedicate this manuscript to all essential workers during these troubling times. “The best way to find yourself is to lose yourself in the service of others.”- Mahatma Gandhi

